# The Impact of Body Mass Index on Robotic Surgery Outcomes in Endometrial Cancer

**DOI:** 10.1101/2023.09.27.23296160

**Authors:** Eva Kadoch, Yoav Brezinov, Gabriel Levin, Florentin Racovitan, Susie Lau, Shannon Salvador, Walter H. Gotlieb

**Affiliations:** Lady Davis Institute of Medical Research, Jewish General Hospital, Montreal, QC, Canada; Experimental Medicine, McGill University, Montreal, QC, Canada; Experimental Surgery, McGill University, Montreal, QC, Canada; Division of Gynecologic Oncology, Sir Mortimer B. Davis Jewish General Hospital, McGill University, Montreal, QC, Canada

**Keywords:** Cohort study, endometrial cancer, obesity, robotics

## Abstract

**OBJECTIVES:** To compare surgical outcomes of patients with endometrial cancer who underwent robotic surgery across different BMI categories.

**METHODS:** A retrospective study including all consecutive patients with endometrial cancer who underwent robotic surgery at a tertiary cancer center between December 2007 and December 2022. The study analyzed outcome measures, including blood loss, surgical times, length of hospitalization, perioperative complications, and conversion rates with the Kruskal-Wallis test for BMI group differences and the Chi-squared test for associations between categorical variables.

**RESULTS:** A total of 1,329 patients with endometrial cancer were included in the study. Patients were stratified by BMI: <30.0 (n=576; 43.3%), 30.0-39.9 (n=449; 33.8%), and ≥40.0 (n=304; 22.9%). There were no significant differences in post-anesthesia care unit (PACU) stay (p=0.105) and hospital stay (p=0.497) between the groups. The rate of post-op complications was similar across the groups, ranging from 8.0% to 9.5% (p=0.761). The rate of conversion to laparotomy was also similar across the groups, ranging from 0.7% to 1.0% (p=0.885). Women with a BMI ≥40.0 had a non-clinically relevant but greater median estimated blood loss (30 mL vs. 20 mL; p<0.001) and longer median operating room (OR) time (288 minutes vs. 270 minutes; p<0.001). Within the OR time, the median set-up time was longer for those with a higher BMI (58 minutes vs. 50 minutes; p<0.001). However, skin-to-skin time (209 minutes vs. 203 minutes; p=0.202) and post-op time (14 minutes vs. 13 minutes; p=0.094) were comparable between groups.

**CONCLUSION:** BMI does not affect the peri-operative outcome of patients undergoing robotic staging procedures for endometrial cancer.

## INTRODUCTION

The prevalence of endometrial cancer is on the rise, possibly due to aging populations and higher obesity rates (1). In 2023, endometrial cancer affected over 66,000 women in the United States, making it the most common gynecological malignancy (2). However, patients with endometrial cancer often face challenges as surgical candidates due to the risk factors associated with the disease, such as obesity and medical comorbidities, making them susceptible to surgical complications (3).

Obesity is a well-known underlying cause of cancer and is directly associated with more than half of cancer cases in the United States (4, 5). Endometrial cancer presents a notable correlation with obesity, with a risk ratio of 1.52 (6-8). Studies consistently show that a significant number of women with endometrial cancer are overweight or obese, underscoring the value to expand our understanding of this relationship (8).

The impact of obesity extends beyond disease incidence, as it may also influence surgical outcomes in gynecological malignancies. Women who are obese may experience larger tumor size, longer surgical times, and increased risk of postoperative complications (6, 9). Moreover, inducing anesthesia in patients with obesity poses challenges and often requires an experienced anesthesia team to consider the anatomic and physiologic changes associated with obesity, including cardiovascular disease and obstructive sleep apnea syndrome (10, 11).

Despite the growing body of research, there remains a lack of large cohort studies investigating the impact of obesity on surgical outcomes in gynecological malignancies. Our study aims to investigate how obesity affects the outcomes of patients undergoing robotic surgery. By conducting a large cohort study on this subject, we aim to provide a comprehensive and up-to-date assessment of the effects of obesity on various aspects of surgical outcomes.

## METHODS

### Study Design

This is a retrospective cohort study of all consecutive women with a preoperative diagnosis of endometrial cancer, who underwent robotic surgery between December 2007 and December 2022, at the Jewish General Hospital in Montreal, Canada. The patients were stratified into three BMI groups based on established criteria for obesity. The non-obese group included patients with a BMI less than 30.0 kg/m^2^, the obese group included patients with a BMI ranging from 30.0 to 39.9 kg/m^2^, and the morbidly obese group included patients with a BMI of 40.0 kg/m^2^ or higher. These cutoffs were selected based on widely accepted classifications for obesity in clinical practice (12-14).

### Ethical Approval

The Research Ethics Committee of the Jewish General Hospital gave ethical approval for this work (2019-1292, 03-041).

### Operative Procedure

The Division of Gynecologic-Oncology at the Jewish General Hospital uses the da Vinci Surgical System since December 2007 for performing robotic surgeries on patients with endometrial cancer, including total hysterectomy, bilateral salpingo-oophorectomy, peritoneal washings, and lymph node evaluation (9). Lymph node evaluation included standard lymphadenectomy until 2010, followed by the sentinel lymph node protocol since. During surgery, patients are continuously monitored with an arterial line, have pneumatic compression stockings, receive antibioprophylaxis with cefazolin, and thromboprophylaxis with subcutaneous heparin. Patients are maintained in a steep Trendelenburg position (25-30 degrees) with insufflation pressures mostly between 10-15 mmHg.

### Data Collection

All data was extracted from the Jewish General Hospital’s electronic medical records system. Baseline characteristics collected included age, BMI, and American Society of Anesthesiologists (ASA) score. Pertinent surgical outcomes such as procedure time, operating room (OR) time, estimated blood loss (EBL), rate of conversion to laparotomy, length of post-anesthesia care unit (PACU) stay, length of hospitalization, and perioperative complications were also collected up to 30 days postop.

### Statistical Analysis

Statistical analyses were carried out using IBM SPSS Statistics, version 28.0 (IBM Corp, Armonk, NY, USA). The normality of the data was assessed using the Shapiro-Wilk test prior to conducting the analyses. Nonparametric tests, specifically the Kruskal-Wallis test, as well as one-way ANOVA followed by a Games-Howell post-hoc comparison test, were used as appropriate. Pairwise comparisons were performed to examine specific differences between BMI groups. To account for multiple comparisons, significance values were adjusted using the Bonferroni correction method. The Pearson’s chi-squared test was used to evaluate associations between categorical variables. A p-value of 0.05 was considered statistically significant. Descriptive parameters were presented as median and interquartile range (IQR). Frequencies were presented as percentages.

### Definitions

Non-obese is defined as a BMI <30.0 kg/m^2^, obese is defined as a BMI of 30.0-39.9 kg/m^2^, and morbidly obese is defined as a BMI of 40 kg/m^2^ or greater (15). We defined OR time as the interval from the patient’s arrival in the OR to their exit. Set-up time was defined as the interval from the patient’s arrival in the OR to the first incision, and skin-to-skin time (also referred to as procedure time) as the interval from the first incision to skin closure. Post-op time refers to the interval from skin closure to the patient’s exit from OR. PACU time denotes the interval from OR exit to discharge from the PACU unit, while in-patient time refers to the interval from PACU unit discharge to hospital discharge.

## RESULTS

### Participant Demographics and BMI Stratification

The studied cohort consisted of 1,329 women with BMI ranging from 16.5 to 85.6 kg/m^2^. BMI were classified into three categories: <30.0 (n=576; 43.3%), 30.0-39.9 (n=449; 33.8%), and ≥40.0 (n=304; 22.9%). The participants had an overall median (IQR) BMI of 31.3 (12.8) kg/m^2^ and overall age of 64.0 (16.0) years (Table 1). Basic patient demographics are presented in Table 1, and the BMI distribution is illustrated in Figure 1.

**Table 1:**
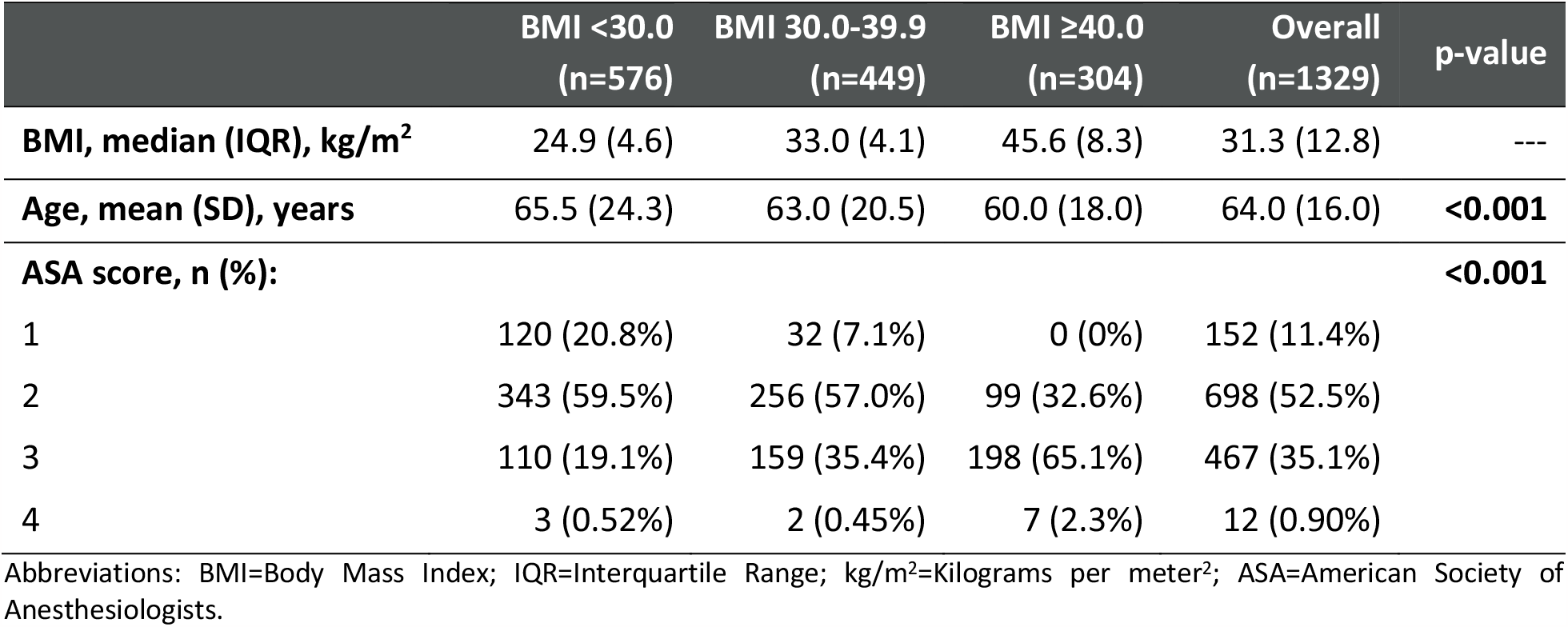
Baseline Demographics and Clinical Characteristics of Endometrial Cancer Patients Undergoing Robotic Surgery.

|  | BMI <30.0<br>(n=576) | BMI 30.0-39.9<br>(n=449) | BMI ≥40.0<br>(n=304) | Overall<br>(n=1329) | p-value |
| --- | --- | --- | --- | --- | --- |
| **BMI, median (IQR), kg/m<sup>2</sup>** | 24.9 (4.6) | 33.0 (4.1) | 45.6 (8.3) | 31.3 (12.8) | --- |
| **Age, mean (SD), years** | 65.5 (24.3) | 63.0 (20.5) | 60.0 (18.0) | 64.0 (16.0) | **<0.001** |
| **ASA score, n (%):** |  |  |  |  | **<0.001** |
| 1 | 120 (20.8%) | 32 (7.1%) | 0 (0%) | 152 (11.4%) |  |
| 2 | 343 (59.5%) | 256 (57.0%) | 99 (32.6%) | 698 (52.5%) |  |
| 3 | 110 (19.1%) | 159 (35.4%) | 198 (65.1%) | 467 (35.1%) |  |
| 4 | 3 (0.52%) | 2 (0.45%) | 7 (2.3%) | 12 (0.90%) |  |

Abbreviations: BMI=Body Mass Index; IQR=Interquartile Range; kg/m<sup>2</sup>=Kilograms per meter<sup>2</sup>; ASA=American Society of Anesthesiologists.

**Figure 1.**
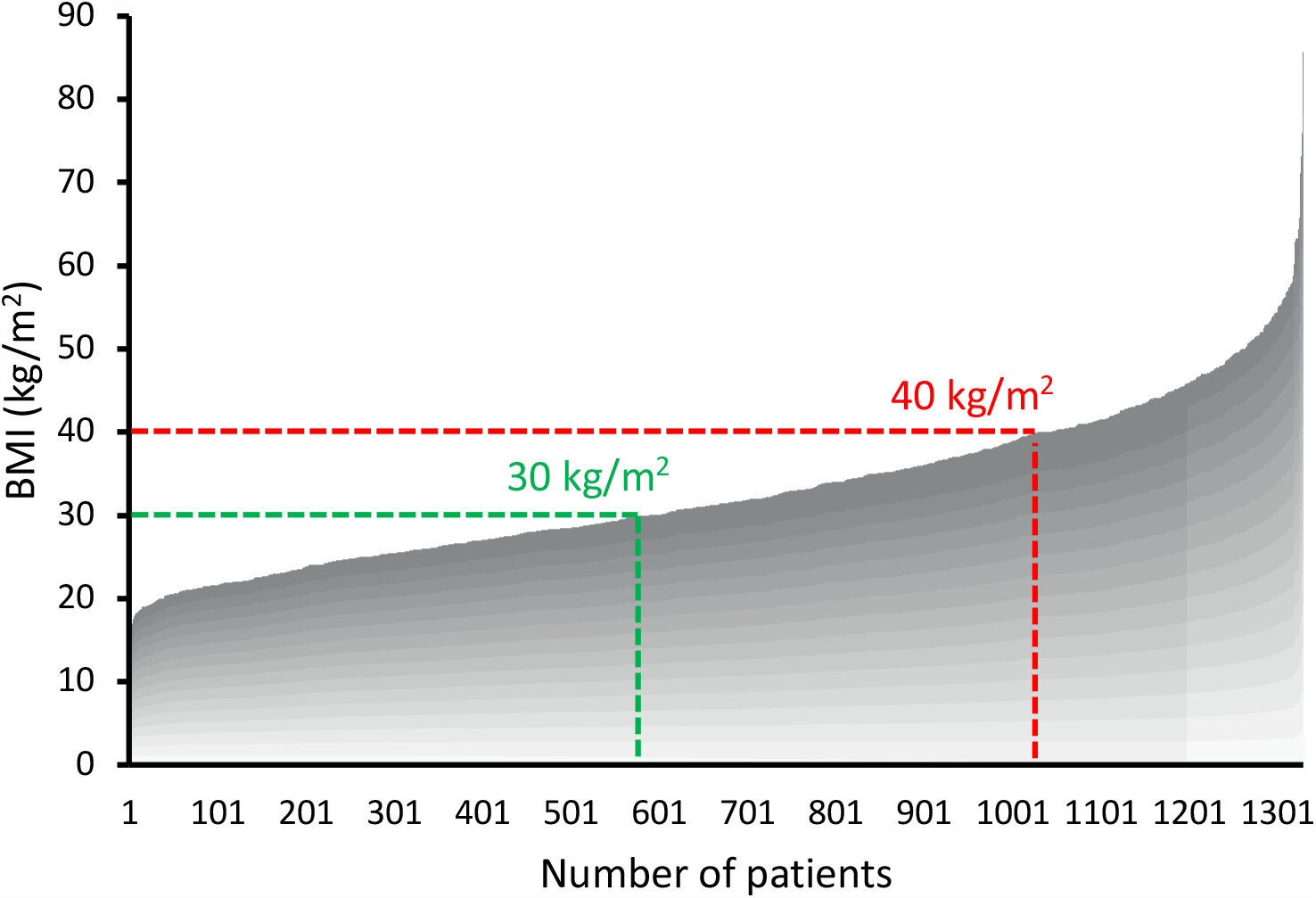
Distribution of BMI in a population of 1,329 endometrial cancer patients.

### Comparison of Surgical Times and Outcomes Among BMI Categories

Women with a BMI ≥40.0 had a longer OR time compared to the other groups (288 minutes vs. 270 minutes: p<0.001) (Figure 3). Within the OR time, the median set-up time was comparable between nonobese and obese groups (50 minutes vs. 51 minutes, respectively), whereas in the morbidly obese group, it was significantly longer (58 minutes; p<0.001) (Figure 2). Median skin-to-skin and post-op times were similar across the groups, ranging between 195 and 209 minutes for skin-to-skin (p=0.202) and 13 and 14 minutes for post-op (p=0.094) (Figure 2). Patients in the morbidly obese group had significantly greater but clinically not relevant EBL (30.0 mL) compared to the non-obese and obese groups (20.0 mL; p<0.001) (Table 2). The median length of hospitalization remained unchanged across the three different groups, with a duration of 1 day in the nonobese group, in the obese group, and in the morbidly obese group (p=0.497) (Table 2). As for the PACU stay, the median was 178 minutes for the nonobese group, 173 minutes for the obese group, and 192 minutes for the morbidly obese group (p=0.105) (Figure 3).

**Table 2:** Surgical Outcomes of Endometrial Cancer Patients Who Underwent Robotic Surgery.

|  | BMI <30.0<br>(n=576) | BMI 30.0-39.9<br>(n=449) | BMI ≥40.0<br>(n=304) | p-value |
| --- | --- | --- | --- | --- |
| EBL, median (IQR), mL | 20 (40) | 20 (37.5) | 30 (35) | <0.001 |
| Hospitalization, median (range), d | 1 (0-12) | 1 (0-6) | 1 (0-7) | 0.497 |
| Conversion to laparotomy, (%) | 0.9 | 0.7 | 1.0 | 0.885 |
| Intra-op complication, (%) | 13.6 | 17.0 | 18.6 | 0.179 |
| ER Presentation, (%) | 8.9 | 8.0 | 9.5 | 0.761 |

Abbreviations: BMI=Body Mass Index; IQR=Interquartile Range; EBL=Estimated Blood Loss; mL=millimeter; ER=Emergency Room.

**Figure 2.**
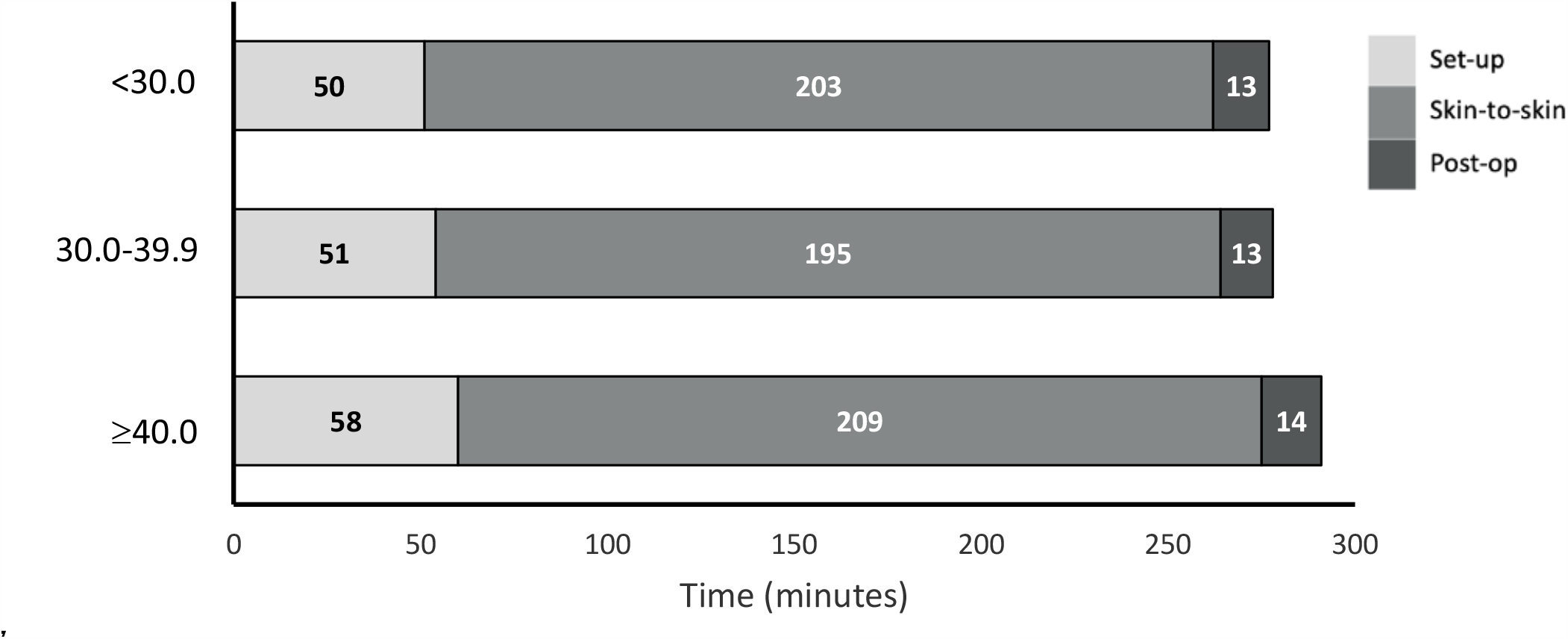
Comparison of OR time components across BMI categories. Abbreviations: Post-op=Post Operation.

**Figure 3.**
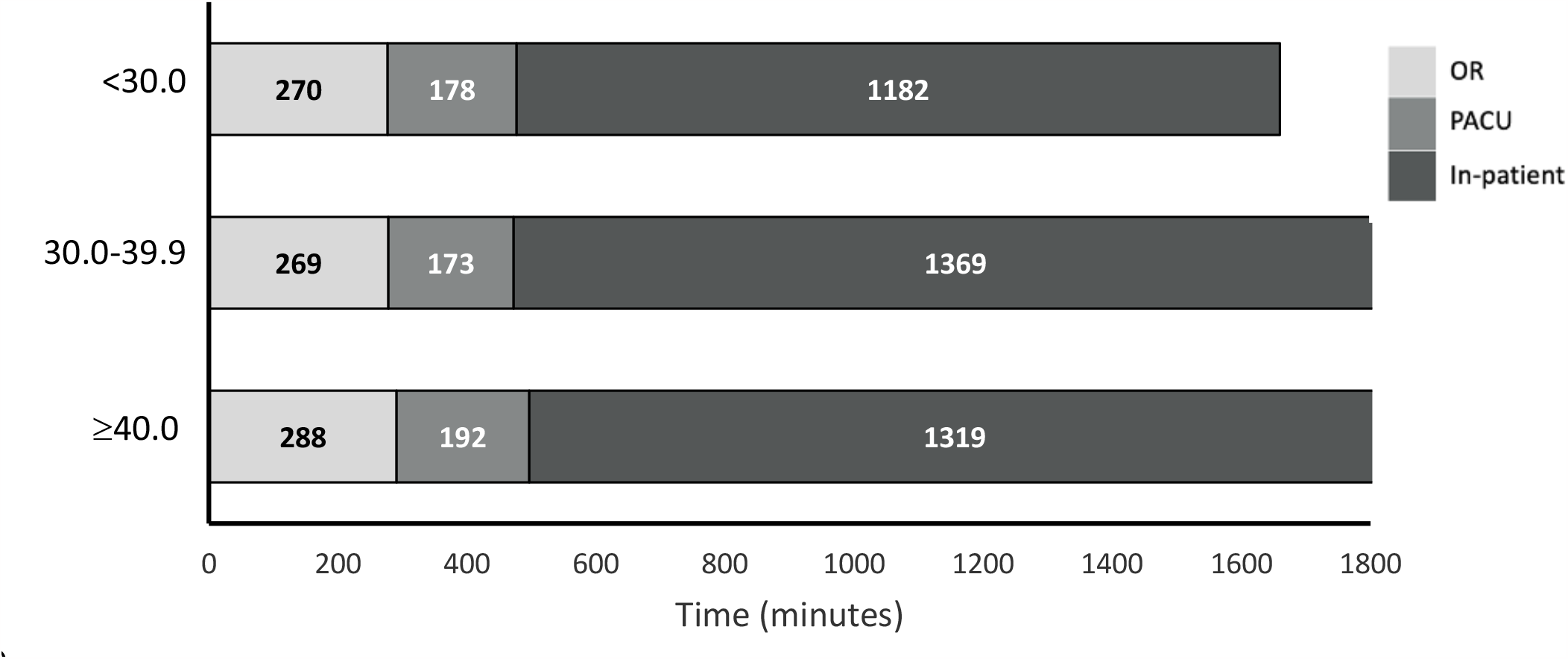
Comparison of post-OR time components across BMI categories. Abbreviations: OR=Operating Room; PACU=Post-Anesthesia Care Unit.

### Complications Rates Among BMI Categories

Intra-operative complications occurred at a similar rate across BMI categories, ranging from 13.6% to 18.6% (p=0.179) (Table 2), with vaginal laceration being the most common intra-operative complication among all BMI groups (Figure 4). In terms of post-operative complications, the BMI ≥40.0 group had the highest rate of emergency room (ER) presentations within 30-days post-op at 9.5%, compared to 8.9% and 8.0% in the other BMI groups (p=0.761) (Table 2). Up to 5% of ER presentations across all BMI groups were for pain and discomfort (Figure 5). Table 1 also shows that conversion rates to laparotomy were low across all BMI groups, remaining below 1.0% (p=0.885).

**Figure 4.**
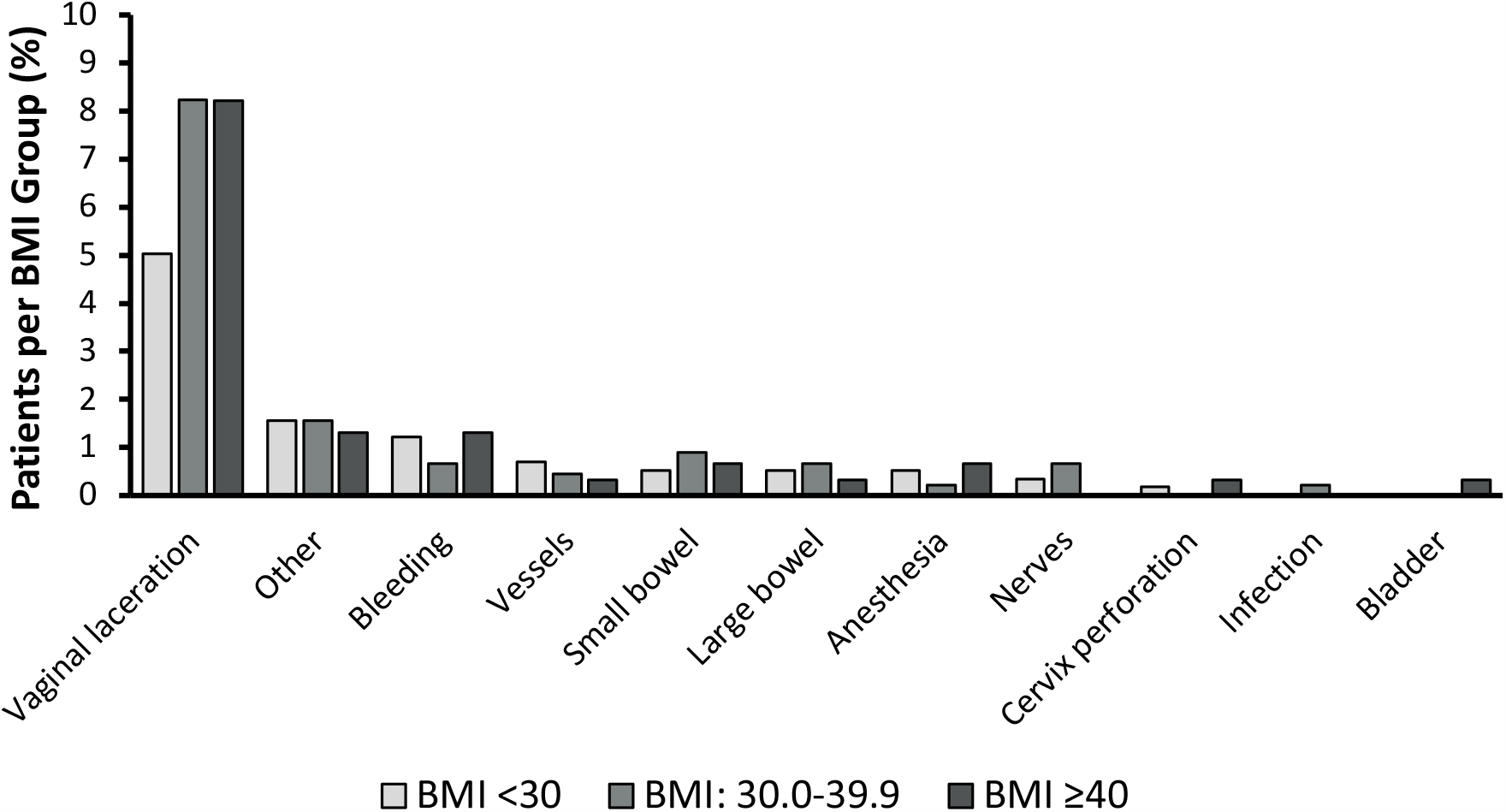
Intra-operative complications.

**Figure 5.**
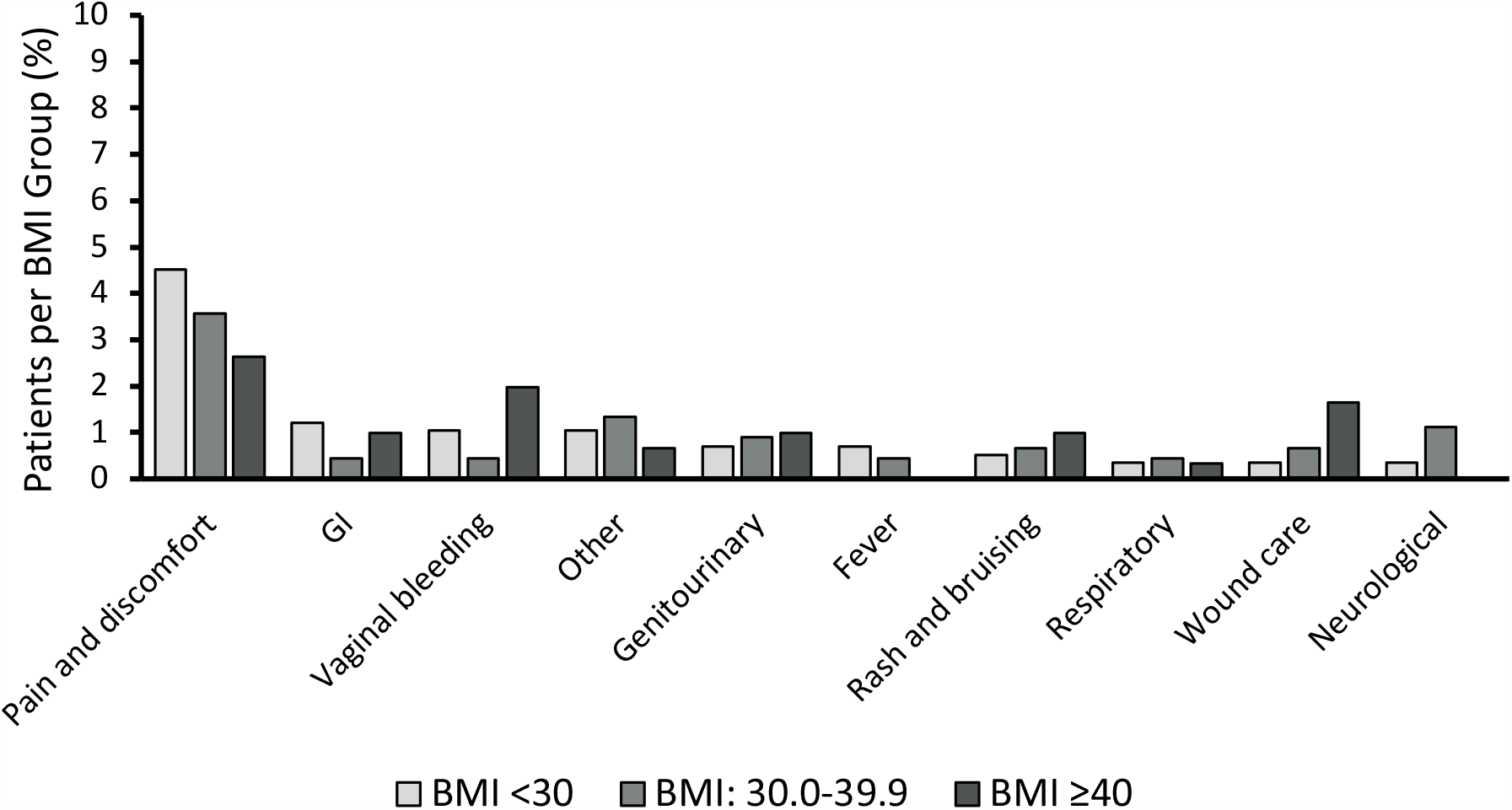
Emergency room presentations 30 days post-op. Abbreviations: GI=Gastrointestinal.

### Descriptive Analysis of Surgical Outcomes in Patients with BMI ≥50.0 kg/m^2^

In addition to the main results described above, a descriptive analysis focusing on patients with a BMI ≥50.0 kg/m^2^ (n=69; 5.2%) was conducted ad hoc, as it was not part of the original study design. We found that patients with a BMI ≥50.0 had median OR times of 297 minutes, median PACU times of 180 minutes, median EBL of 50 mL, and median length of hospitalization of 1 day. The prevalence of intraoperative complications was 30% in this subgroup with vaginal laceration remaining as the most common complication (17.4%). Two patients (2.9%) underwent a conversion to laparotomy due to intolerable Trendelenburg positioning, and six patients (8.7%) presented to the ER, primarily for wound checks.

## DISCUSSION

In this retrospective study, we examined the relationship between obesity and surgical outcomes in a cohort of 1,329 women with endometrial cancer, with the goal of assessing the safety of robotic surgery among patients who are obese. Despite the increased risk profile associated with obesity, our main findings indicate that surgical outcomes were comparable across the BMI groups.

Understanding how obesity influences surgical outcomes is crucial for patient risk stratification and preoperative planning. Metabolic syndrome, which includes hypertension, dyslipidemia, and type 2 diabetes, is common in women with endometrial cancer and increases the risk of surgical complications (11). Our findings showed that the obese and morbidly obese groups reported significantly higher percentages of individuals with an ASA score ≥2, with 79% of patients for non-obese, 93% for obese, and 100% for morbidly obese (Table 1). Interestingly, surgical outcomes, including length of hospitalization, and PACU stay, peri-operative complication rates, and conversion rates were found to be comparable across the three BMI groups despite this elevated risk profile.

We observed an increase in overall operating room (OR) time in patients with higher BMI, with the morbidly obese group experiencing the most significant difference. This finding is consistent with previous studies that have reported longer surgical times in obese patients undergoing robotic surgery (18, 19). A further analysis of the OR time components revealed that set-up time was significantly longer in the morbidly obese group, whereas skin-to-skin time and post-op times were consistent across BMI categories (Figure 2). This difference in set-up time may be attributed to the challenges associated with patient positioning, anesthesia induction, or trocar placement in patients who are morbidly obese. By identifying specific challenges associated with obesity, the surgical team can implement tailored strategies to mitigate these risks. For instance, optimizing anesthesia protocols can help address the physiological changes associated with obesity, ultimately improving patient safety and minimizing adverse events (10). Overall, intra-operative complications were low, but somewhat higher with increasing BMI, although no significant differences were observed. Minor vaginal lacerations were by far the most common complication, occurring in all BMI groups (Figure 4). This complication is often attributed to the technical challenges of removing larger specimens through the vaginal route. ER presentations were low, with pain and discomfort being the most common reason in all BMI groups (Figure 5). The findings of our study show that patients with elevated BMI can successfully benefit from minimally invasive surgery without significantly increasing their risk.

The primary focus of our study was to examine the relationship between obesity and robotic surgery outcomes within the standard BMI categories: <30, 30.0-39.9, and ≥40.0 kg/m^2^. In addition, we conducted an ad-hoc descriptive analysis on patients with a BMI ≥50.0 kg/m^2^. In this small subgroup of patients, we noted that OR times and EBL were in line with the trends observed in the three original BMI groups. Despite an increased prevalence of complications, mainly vaginal lacerations, the study suggests that patients with a BMI ≥50.0 kg/m^2^ benefit from minimally invasive surgery.

Our study is not without limitations. First, the retrospective nature of this analysis inherently presents a risk for confounding. Second, this study was conducted at a single center, specifically a university hospital which could have resulted in longer surgical times, especially in the non-obese group due to trainees being assigned the less complex cases. Third, the study focuses primarily on short-term surgical outcomes, with limited analysis of long-term outcomes and quality of life. Further research is needed to evaluate the impact of obesity on long-term oncologic outcomes in a large cohort of patients.

In conclusion, the comparable surgical outcomes and complication rates across BMI groups associated with the use of robotic surgery which provides several technological advantages over traditional surgical approaches. Our results support robotic surgery as a feasible option for patients with obesity who were diagnosed with endometrial cancer and can help overcome the difficulties associated with patient obesity during surgery.

## ABBREVIATIONS

ANOVA: Analysis of Variance
ASA: American Society of Anesthesiologists
BMI: Body Mass Index
EBL: Estimated Blood Loss
EIN: Endometrial Intraepithelial Neoplasia
ER: Emergency Room
IQR: Interquartile Range
OR: Operating Room
PACU: Post-Anesthesia Care Unit
SD: Standard Deviation

## AUTHOR CONTRIBUTIONS

WH.G., S.L., S.S. designed the study. E.K., Y.B., G.L. participated in the primary analysis. F.R. and E.K. participated in data collection. E.K. analyzed the data and wrote the manuscript. All authors discussed the results, provided critical input, edited the manuscript, and provided final approval of the version to be published. All authors agree to assuming accountability for all aspects of the study, thereby ensuring thorough investigation and resolution of any inquiries concerning the accuracy or integrity of the work.

## Data Availability Statement

All data produced in the present study are available upon reasonable request to the authors.

### Approvals

The study was approved by the Research Ethics Board of the Jewish General Hospital (2019-1292, 03-041).

## Declaration of Interests

The authors have no conflicts of interest to disclose.

### Declaration of AI-Assisted Technologies in the Writing Process

During the preparation of this work, the authors used ChatGPT in order to improve language and sparingly assist in the translation of text to English. After using this tool, the authors reviewed and edited the content as needed and take full responsibility for the content of the publication.

